# Predicting arterial age using carotid ultrasound images, pulse wave analysis records, cardiovascular biomarkers and deep learning

**DOI:** 10.1101/2021.06.17.21259120

**Authors:** Alan Le Goallec, Sasha Collin, Samuel Diai, Théo Vincent, Chirag J. Patel

**Affiliations:** Department of Biomedical Informatics, Harvard Medical School, Boston, MA, 02115, USA; Department of Systems, Synthetic and Quantitative Biology, Harvard University, Cambridge, MA, 02118, USA

## Abstract

Cardiovascular disease --an age-related disease-- is the leading cause of death worldwide. We built an arterial age predictor by training deep learning models to predict age from 233,388 pulse wave analysis records, 8,279 carotid ultrasound images and arterial health biomarkers (e.g blood pressure) collected from 502,000 UKB participants. We predicted age with a R-Squared of 67.1+/-0.6% and a root mean squared error of 4.29+/-0.04 years. Attention maps for carotid ultrasound images suggest that the predictions are driven by vascular features, for the largest part. Accelerated arterial aging is 32.6+/-7.3% GWAS-heritable, and we identified 192 single nucleotide polymorphisms in 109 genes (e.g NPR3, involved in blood volume and pressure) significantly associated with this phenotype. Similarly, we identified biomarkers (e.g electrocardiogram features), clinical phenotypes (e.g chest pain), diseases (e.g hypertension), environmental (e.g smoking) and socioeconomic (e.g income and education) variables associated with accelerated arterial aging. Finally, carotid ultrasound images, pulse wave analysis records and blood pressure biomarkers capture different facets of arterial aging. For example, carotid ultrasound-measured and pulse wave analysis-measured accelerated arterial aging phenotypes are only .164+/-.009 correlated. In conclusion, our predictor suggests potential lifestyle and therapeutic interventions to slow arterial aging, and could be used to assess the efficiency of emerging rejuvenating therapies on the arterial system.

## Introduction

Cardiovascular disease [CVD] is the leading cause of death worldwide with 17.8 millions of deaths every year ^1^. This corresponds to 330 millions years of life lost ^2^ and 36 millions years of life with disability ^3^. With the world population aging and CVD being a chronic disease with age as a major risk factor, its prevalence is expected to increase. Understanding the aging of the cardiovascular system could therefore be key in reducing the burden of and treating CVD diseases.

Here, we studied arterial aging using the concept of “biological age”. In contrast to chronological age, the time elapsed since an individual’s birth, biological age represents the effects of time on the participants body, and is the true underlying cause of age-related diseases such as CVD. “Accelerated agers” are individuals whose biological age is higher than their chronological age. Biological age predictors can be built by training a machine learning model (e.g linear regression, gradient boosted machine, neural network) to predict chronological age from physiological measures (e.g blood pressure). After the algorithm has been trained, the predictions it outputs for unseen data can be interpreted as the biological age of the participant.

Measures of arterial health such as blood pressure and arterial thickness represent promising candidate biomarkers to build arterial age predictors, because the vascular system undergoes important changes with age. For example, atherosclerosis develops, so intima media thickness increases ^4^ and arterial vessels stiffen, which leads to an increase in systolic blood pressure and decrease in diastolic blood pressure ^5^. Fedintsev et al. predicted chronological age from scalar features extracted from carotid artery duplex scan and pulse wave velocimetry (R-Squared [R^2^]=55-69%; root mean squared error [RMSE]=5.87-6.91 years) ^6^. Cardiac features such as electrocardiograms ^7^ and heart MRI videos ^8^ have similarly been used to build chronological age predictors. Framingham’s ^9^ heart age uses a conceptually different approach, directly adjusting chronological age for factors such as smoking, blood pressure and cholesterol ^10^.

A key unanswered question is whether additional information regarding the arterial aging process can be uncovered by leveraging raw carotid ultrasound images and pulse wave analysis records, as opposed to relying on summary scalars features extracted from the aforementioned datasets. Additionally, the genetic and non-genetic factors of accelerated arterial aging remain elusive.

In this paper, we leveraged 235,000 pulse wave analysis records (Figure 1D), 9,000 carotid ultrasound images (Figure 1C) and arterial health biomarkers (e.g blood pressure) collected from 502,000 UKB participants aged 45-81 years to predict chronological age (Figure 1A and B). We then performed a genome wide association study [GWAS] to estimate the heritability of accelerated arterial aging and identify single nucleotide polymorphisms [SNPs] associated with this phenotype. Similarly, we performed an X-wide association study [XWAS] to identify biomarkers, clinical phenotypes, diseases, family history, environmental and socioeconomic variables associated with accelerated arterial aging. (Figure 1E)

**Figure 1:**
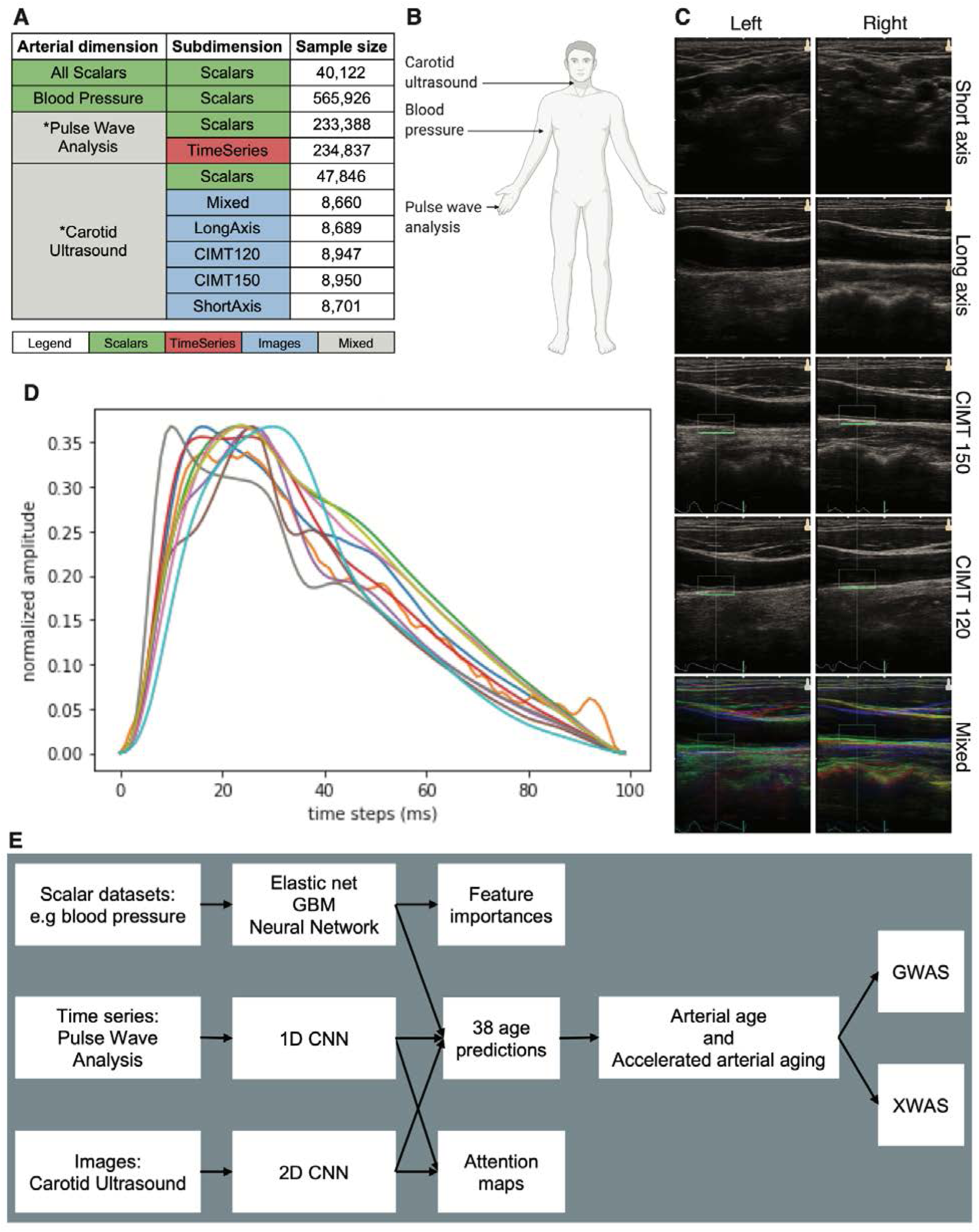
Datasets and pipeline overview. A - Arterial aging measurement modalities. B - Body sites where arterial health data was collected. C - Carotid ultrasound image samples. D - Ten pulse wave analysis samples. E - Overview of the analytical pipeline.

## Results

### Leveraging raw images and times series with deep learning improved age prediction performance

We performed our analysis (Figure 1E) on 502,000 UK Biobank participants aged 45-81 years (Fig. S1). Specifically, we predicted chronological age from 8,279 carotid ultrasound images (R^2^=64.8+/-1.9%; RMSE=4.44+/-0.13 years), 233,338 pulse wave analysis records (R^2^=41.3+/-0.3%; RMSE=6.53+/-0.04 years) and 565,926 blood pressure biomarkers samples (R^2^=22.3+/-0.3%; RMSE=7.34+/-0.02 years). We ensembled these models and predicted age with a R^2^ value of 67.1+/-2.0% and a RMSE=4.29+/-0.13 years. (Figure 2)

**Figure 2:**
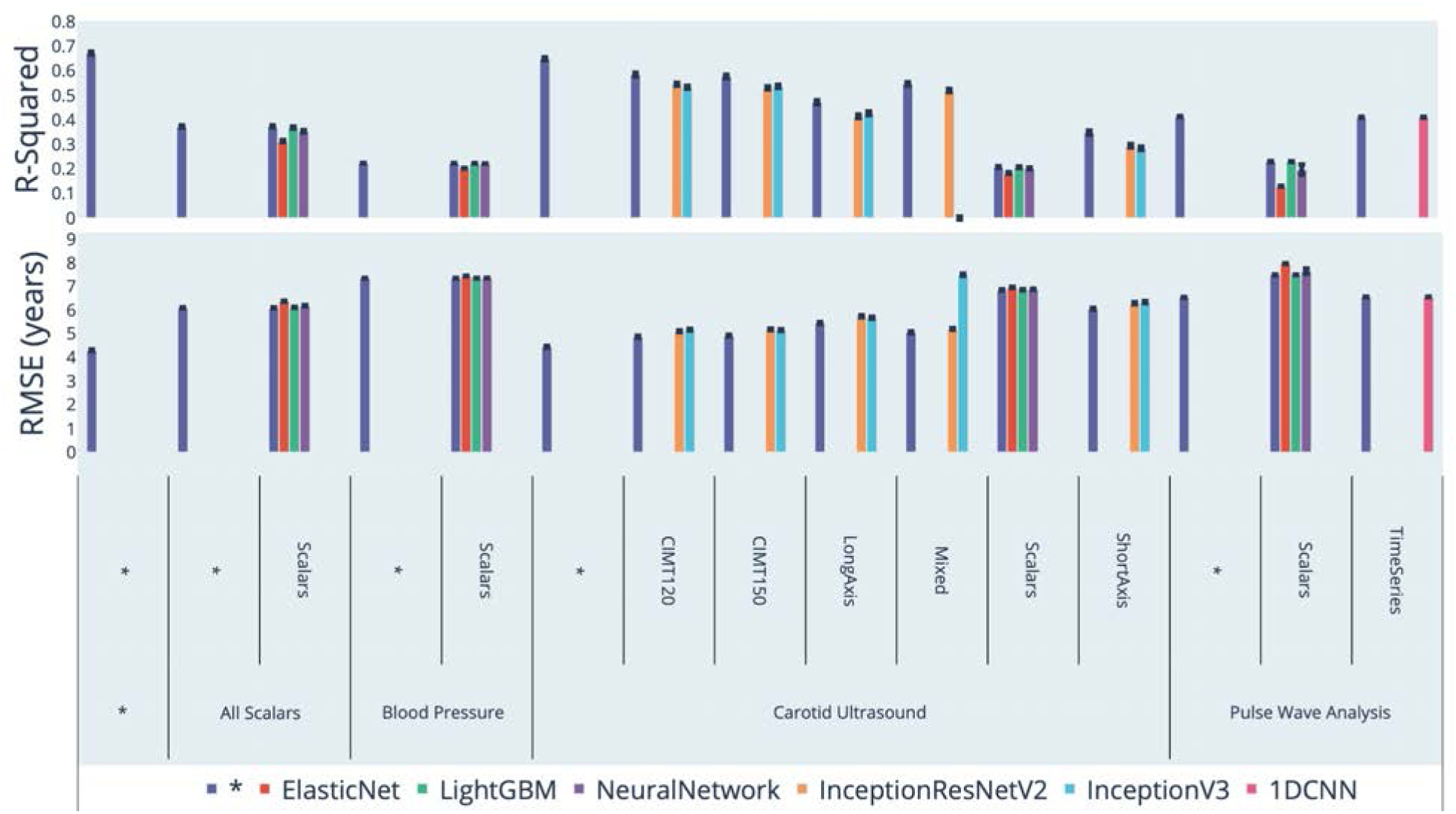
Chronological age prediction performance from carotid ultrasound images, pulse wave analysis records and blood pressure measurements All statistics computed on the testing set. * represent ensemble models

The models built on the raw data significantly outperformed the models built on the scalar features (Table 1). Specifically, the model built on carotid ultrasound images explained three times more chronological age variance than the model built on carotid ultrasound scalar features (64.8+/-1.9% vs. 20.7+/-1.6%). The model built on raw pulse wave analysis records explained approximately twice more chronological age variance than the model built on scalar predictors (41.3+/-0.3% vs. 23.0+/-0.7%). Finally, the ensemble model encompassing all datasets increased the R^2^ by 30% compared to the ensemble model limited to scalar predictors (67.1+/-2.0% vs. 37.2+/-1.5%).

**Table 1:** Comparison between the prediction performances of raw data-based and scalar features-based age predictors

| Dataset | R-Squared (%) |  | RMSE (years) |  |
| --- | --- | --- | --- | --- |
|  | Raw data | Scalar features | Raw data | Scalar features |
| Carotid ultrasound | 64.8±1.9 | 20.7±1.6 | 4.44±0.13 | 6.85±0.7 |
| Pulse wave analysis | 41.3±0.3 | 23.0±0.7 | 6.53±0.04 | 7.48±0.05 |
| Blood pressure | NA | 22.3±0.3 | NA | 7.34±0.02 |
| Ensemble | 67.1±2.0 | 37.2±1.5 | 4.29±0.13 | 6.09±0.08 |

We report the performance and number of predictors for each model (Table S6), as well as the Pearson and Spearman correlation between the feature importance for each algorithm (Table S7 and Table S8).

### Identification of the features driving arterial age prediction

For the models built on carotid ultrasound images, our attention maps highlighted the carotid as the primary feature driving the chronological age prediction. Other anatomical features of interest to the model included the surrounding tissue, jugular veins and the skin tissue (Figure 3). Similarly, we used attention maps on pulse wave analysis records (Fig. S2). Additional attention maps can be found at https://www.multidimensionality-of-aging.net/ under “Model Interpretability”.

**Figure 3:**
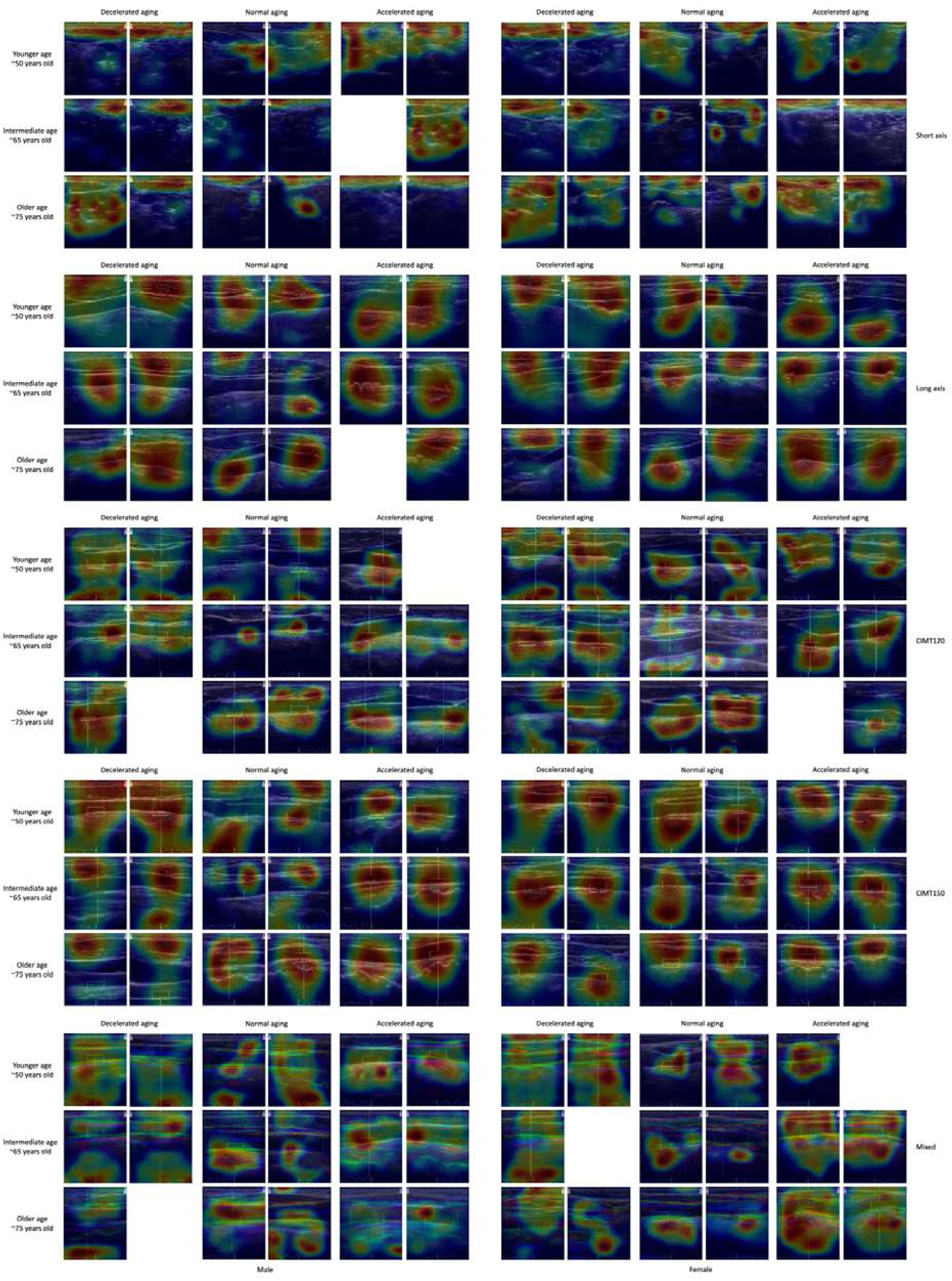
Sample carotid ultrasound images attention maps

For the models built on scalar features, the best performing non-ensemble model (a gradient boosted machine, R^2^=36.8+/-0.4%) reported the following ten features as most important. (1) pulse wave peak to peak time, (2) pulse wave arterial stiffness index, (3) systolic blood pressure, (4) diastolic blood pressure, (5) pulse rate, (6) pulse wave reflection index, (7) mean carotid intima-medial thickness (IMT) at 120 degrees, (8) mean carotid intima-medial thickness (IMT) at 240 degrees, (9) position of pulse wave notch and (10) mean carotid intima-medial thickness (IMT) at 210 degrees. For the exhaustive list and feature importance values for the different models, please refer to Fig. S3, Table S2, Fig. S4, Table S3, Fig. S5, Table S4, Fig. S6, Table S5.

### Genetic factors and heritability of accelerated arterial aging

We defined accelerated arterial aging for each sample as the difference between the predicted age (the participant’s biological age) and chronological age. We performed a genome wide association study [GWASs] and found this phenotype to be 32.6+/-7.3% GWAS-heritable. We performed a similar analysis on accelerated arterial aging as solely defined by pulse wave analysis, and as solely defined by carotid ultrasound images. We found these two phenotypes to be respectively 17.2+/-0.4% and 28.6+/-7.0% GWAS-heritable.

We identified 192 SNPs in 109 genes significantly associated with pulse wave analysis-based accelerated arterial aging (Figure 4). The ten highest GWAS peaks are: (1) AC073115.6 (a long intergenic non-coding RNA). AC073115.6 is in linkage disequilibrium with CDK6 (Cyclin Dependent Kinase 6, involved in cell cycle regulation); (2) CNNM2 (Cyclin and CBS Domain Divalent Metal Cation Transport Mediator 2, involved in magnesium homeostasis); (3) IGFBP3 (Insulin Like Growth Factor Binding Protein 3, a growth factor regulator); (4) TEX41 (a long non-coding RNA linked to aortic valve disease 1); (5) HMGA2 (High Mobility Group AT-Hook 2, a transcription regulator); (6) ATP1B3 (ATPase Na+/K+ Transporting Subunit Beta 3, involved in maintaining Na and K ions’ electrochemical gradients and in cell-surface interactions at the vascular wall); (7) MTND3P1 (Mitochondrially Encoded NADH:Ubiquinone Oxidoreductase Core Subunit 3, a pseudogene); (8) NPR3 (Natriuretic Peptide Receptor 3, involved in blood volume and pressure and linked to hypertension); (9) LINC00521 (Coiled-Coil Domain Containing 197 a protein coding gene with unknown function); and (10) CNBD2 (Cyclic Nucleotide Binding Domain Containing 2, involved in cyclic AMP binding and important to male fertility). ^11^ Due to the limited sample size of the ensemble model (N=7,549) and the carotid ultrasound-based model (N=7,885), we did not detect any single nucleotide polymorphisms associated with accelerated arterial aging for these two models (Fig. S7 and Fig. S8).

**Figure 4:**
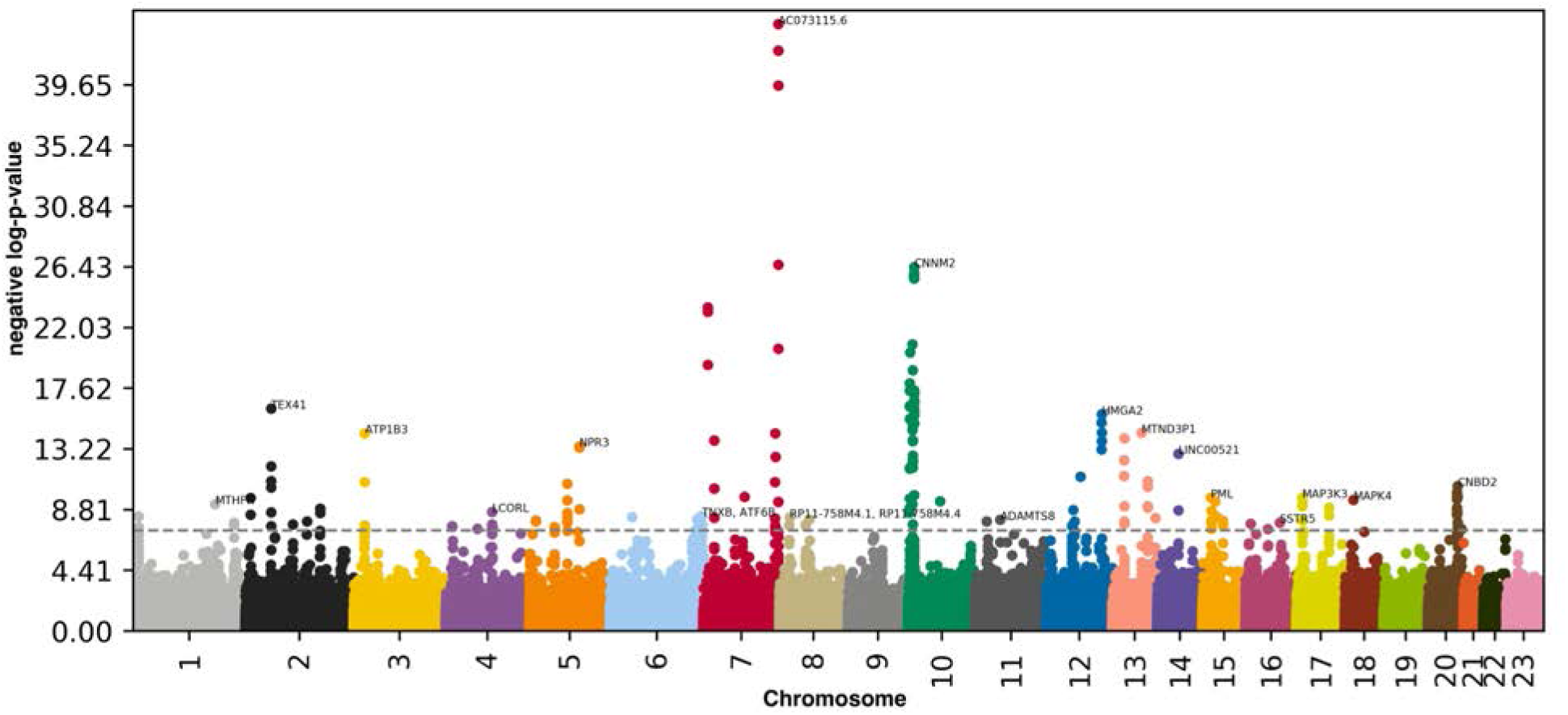
Genome Wide Association Study results for pulse wave analysis-measured accelerated arterial aging negative log10(p-value) vs. chromosomal position. Dotted line denotes 5e-8.

A summary of our GWAS findings can be found in Table S1.

### Non-genetic factors associated with accelerated arterial aging

We performed X-wide association studies [XWASs] to identify biomarkers (Table S9), clinical phenotypes (Table S12), diseases (Table S15), environmental (Table S21) and socioeconomic (Table S24) variables associated with accelerated arterial aging. Please refer to the supplementary tables (Table S10, Table S11, Table S13, Table S14, Table S16, Table S17, Table S19, Table S20, Table S22, Table S23, Table S25, Table S26) for a more detailed summary of non-genetic factors associated with the different facets of accelerated arterial aging (general, pulse wave analysis-based, and carotid ultrasound-based). The full results can be exhaustively explored at https://www.multidimensionality-of-aging.net/xwas/univariate_associations.

#### Biomarkers associated with accelerated arterial aging

The three biomarker categories most associated with accelerated arterial aging, aside from arterial biomarkers categories (carotid ultrasound, pulse wave analysis, blood pressure), are electrocardiogram, anthropometry and blood count (Table S10). Specifically, 25.0% of electrocardiogram biomarkers are associated with accelerated arterial aging, with the three largest associations being with QTC interval (correlation=.072), ventricular rate (correlation=.061), and QRS num (correlation=.060). 25.0% of anthropometry biomarkers are associated with accelerated arterial aging, with the two associations being with waist circumference (correlation=.084) and body mass index (correlation=.077). 22.6% of blood count biomarkers are associated with accelerated arterial aging, with the three largest associations being with neutrophil count (correlation=.078), immature reticulocyte fraction (correlation=.068), and platelet crit (correlation=.067).

Conversely, the three biomarker categories most associated with decelerated arterial aging are spirometry, cognitive symbol digit substitution and hand grip strength (Table S11). Specifically, 100.0% of spirometry biomarkers are associated with decelerated arterial aging, with the three associations being with forced expiratory volume in one second (correlation=.106), forced vital capacity (correlation=.100), and peak expiratory flow (correlation=.065). 100.0% of cognitive symbol digit substitution biomarkers are associated with decelerated arterial aging, with the two associations being with number of symbol digit matches made correctly (correlation=.064) and number of symbol digit matches attempted (correlation=.062). 100.0%% of hand grip strength biomarkers are associated with decelerated arterial aging, with the two associations being with left hand grip strength (correlation=.088) and right hand grip strength (correlation=.088).

#### Clinical phenotypes associated with accelerated arterial aging

Two clinical phenotype categories are associated with accelerated arterial aging: breathing and general health (Table S13). Specifically, 50.0% of breathing clinical phenotypes are associated with accelerated arterial aging, with the association being with the presence of a wheeze or whistling in the chest in the last year (correlation=.056). 25.0% of general health clinical phenotypes are associated with accelerated arterial aging, with the two associations being with overall health rating (correlation=.090) and long-standing illness, disability or infirmity (correlation=.061).

Conversely, the only clinical phenotype category associated with decelerated arterial aging is sexual factors (age first had sexual intercourse: correlation=.067, Table S14).

#### Diseases associated with accelerated arterial aging

Accelerated arterial aging is associated with two diseases: hypertension (correlation=.097) and mental and behavioral disorders due to use of tobacco (correlation=.051). (Table S16)

#### Environmental variables associated with accelerated arterial aging

The three environmental variable categories most associated with accelerated arterial aging are smoking, sun exposure and alcohol intake (Table S22). Specifically, 29.2% of smoking variables are associated with accelerated arterial aging, with the three largest associations being with pack years adult smoking as proportion of life span exposed to smoking (correlation=.107), pack years of smoking (correlation=.103), and smoking status: current (correlation=.072). 10.0% of sun exposure variables are associated with accelerated arterial aging, with the two associations being with time spent outdoors in summer (correlation=.073) and time spent outdoors in winter (correlation=.055). 6.9% of alcohol variables are associated with accelerated arterial aging, with the two associations being with average weekly beer plus cider intake (correlation=.098) and average weekly spirits intake (correlation=.052).

Conversely, the three environmental variable categories most associated with decelerated arterial aging are smoking, medication and physical activity (Table S23). Specifically, 25.0% of smoking variables are associated with decelerated arterial aging, with the three largest associations being with smoking status: never (correlation=.073), current tobacco smoking: no (correlation=.073), and age started smoking in current smokers (correlation=.065). 6.1% of medication variables are associated with decelerated arterial aging, with the two associations being with not taking any medication for cholesterol, blood pressure or diabetes (correlation=.095) and not taking any medication for cholesterol, blood pressure, diabetes or exogenous hormones (correlation=.084). 5.7% of physical activity variables are associated with decelerated arterial aging, with the two associations being with frequency of strenuous sports in the last four weeks (correlation=.055) and duration of strenuous sports (correlation=.053).

#### Socioeconomic variables associated with accelerated arterial aging

The two socioeconomic variable categories associated with decelerated arterial aging are education (college or university degree: correlation=.079) and household (average total household income before bax: correlation=.064). (Table S26)

### Comparison between carotid ultrasound-measured and pulse wave analysis-measured accelerated arterial aging

We computed the correlation between accelerated arterial aging as defined by carotid ultrasound images versus as defined by pulse wave analysis and found that the two are .164+/-.009 correlated (Figure 5). We also used the XWAS results to compute the correlation between these two phenotypes in terms of association with non-genetic variables. We found that, in general, non-genetic variables associated with carotid ultrasound image-measured accelerated arterial aging tend to be similarly associated with pulse wave analysis-measured accelerated arterial aging (Fig. S9).

**Figure 5:**
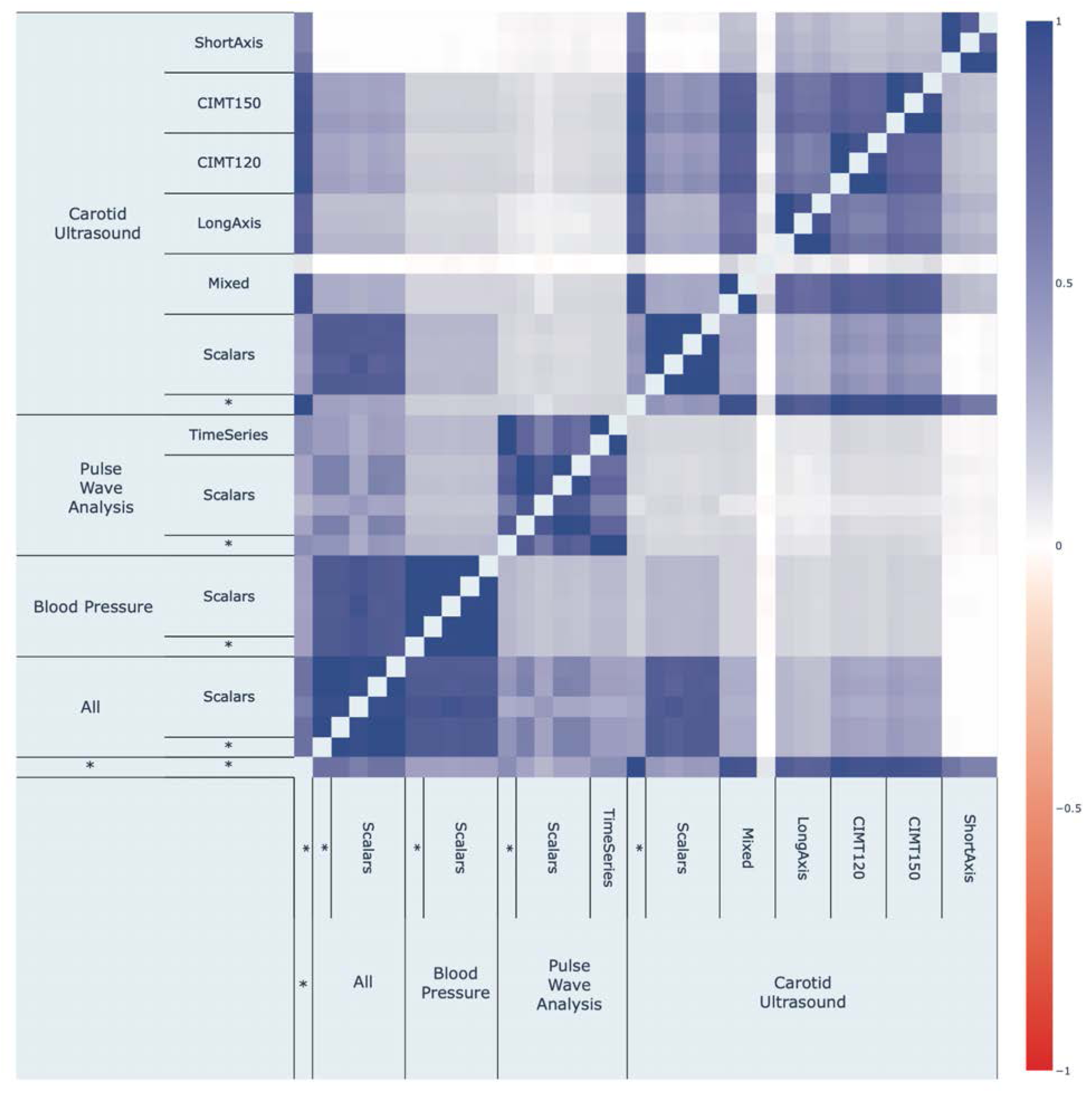
Correlation between accelerated arterial aging as defined by different models

### Prediction of survival from pulse wave analysis

Biological age predictors can be built by training machine learning algorithms to predict an age-related phenotype such as chronological age from medical datasets. Chronological age is a common target, but survival prediction is also a suitable choice. We predicted survival from pulse wave analysis data and obtained a concordance index [CI] of 71.4+/-0.9%, to be compared to a CI of 69.5+/-0.9% for the model trained on the sole demographic features (Fig. S10 and Table S27).

## Discussion

We predicted chronological age with a R^2^ of 67.1+/-0.6% from carotid ultrasounds, pulse wave analysis records and blood pressure biomarkers. Age is best predicted by carotid ultrasound images (R^2^=64.8+/-1.9%), despite their limited sample size (N=8,279). For both carotid ultrasounds and pulse wave analysis records, we found that the models built on the raw data significantly outperformed the models built on scalar features, suggesting that some key information was not summarized by the scalar features extracted from the images and time series by UKB. We therefore recommend that future research and clinical applications of carotid ultrasound images and pulse wave analysis records use convolutional neural networks to extract as much information as possible from the raw data.

Non-linear models (GBM, neural networks) only marginally outperformed linear models (elastic net) when trained on blood pressure or carotid ultrasound-derived scalar features. In contrast, the GBM trained on pulse wave analysis scalar features explained approximately twice more age variance than the elastic net (R^2^=22.9+/-0.3% vs. 12.9+/-0.3%), suggesting that important information regarding aging is non-linearly encoded in complex interactions between the different pulse wave analysis-derived scalar features.

The attention maps for carotid ultrasound images highlighted the carotid as an important feature leveraged by the model to predict age. Other anatomical features were highlighted too, such as jugular veins, the surrounding tissue and the skin tissue. The latter features are potentially problematic. If prediction accuracy is mostly driven by the tissue surrounding the artery and the skin tissue, what we have designed is not an arterial age predictor, but a tissue age predictor. One way to estimate the contribution of the surrounding and skin tissues to the age prediction is to compare the prediction accuracy of the models trained on short axis-view carotid ultrasound images and the models trained on the long axis-view of these same images. The former images make it more difficult for the model to base its prediction on the arterial features, as observed on the attention maps. The difference in prediction performances (short axis: R^2^=34.7±0.8%; long axis: R^2^=47.1±0.8%) suggests that as much as 75% of carotid ultrasound-based “arterial age” prediction could actually be driven by extra-arterial anatomical features, and could explain the low correlation (.164+/-.009) between carotid ultrasound-measured and pulse wave analysis-measured accelerated arterial aging. This last correlation should however be interpreted while keeping in mind that two models trained on longitudinal views of the carotid artery (long axis and CIMT120) can be as weakly as .438+/-.009 correlated. Considering the important contribution of carotid ultrasound images to the prediction of the ensemble arterial age predictor, the “contamination” of our predictor by non-arterial information suggests that its clinical relevance could be significantly improved by first segmenting the carotid ultrasound images to isolate the arterial features. Despite this limitation, the non-genetic factors associated with accelerated arterial aging as defined by the model trained on carotid ultrasound images imply biological relevance. Carotid ultrasound-measured accelerated arterial aging is associated with blood pressure, pulse wave analysis, heart function and electrocardiogram biomarkers, and hypertension.

The biological relevance of the model built on pulse wave analysis records is confirmed by the SNPs associated with the accelerated arterial aging phenotype derived from it. Out of the ten highest peaks highlighted by the GWAS, four are directly linked to arterial health. Specifically, CNNM2 is linked in magnesium homeostasis, which is involved in hypertension and cardiovascular disease ^12–14^, TEX41 is linked to aortic valve disease, ATP1B3 is involved in cell-surface interactions at the vascular wall, and NPR3 involved in blood volume and pressure and linked to hypertension.

Accelerated arterial aging is also associated with biomarkers from diverse organ systems such as the spirometry, cognitive tests, hand grip strength, anthropometry, blood biochemistry, brain MRI features and blood count biomarkers, in addition to self-reported general health factors such as overall health rating. This suggests that accelerated arterial aging is linked to a general aging process that may be correlated with aging processes in other organs. We explore the connection between accelerated arterial aging and aging of other organs in a separate paper ^15^.

In terms of environmental exposures, accelerated arterial aging is associated with smoking (including maternal smoking around birth), and lack of physical activity, in agreement with the literature ^16, 17^. In terms of alcohol intake, beer, cider and spirits intake is associated with accelerated arterial aging, while usually taking alcohol with meals is associated with decelerated arterial aging, reflecting the complex literature on the topic ^18^. We found cheese intake to be associated with decelerated arterial aging, in accordance with the most recent review on the subject ^19^.

Decelerated arterial aging is associated with education, which is possibly mediated by health literacy ^20^. Similarly, it is associated with higher income, providing access to better healthcare. In the US, the richest 1% live approximately a decade older than their poorest 1% counterparts (14.6±0.2 years longer for males, 10.1±0.2 years for females) ^21^.

Pulse wave analysis records only marginally increased survival prediction performance compared to simply relying on demographics (age, sex and ethnicity) to predict death, as is often the case with survival analysis ^22^.

In conclusion, our arterial health predictors capture different facets of arterial aging, and their association with genetic and non-genetic factors suggest potential lifestyle and therapeutic interventions to slow it down. The predictors could also be used to assess the efficiency of emerging rejuvenating therapies ^23^ on the cardiovascular system. Other biological age predictors such as DNA methylation ^24^ are currently being used to assess general rejuvenation ^25^, but aging is multidimensional ^15, 26^ and a single predictor might not capture the effect of a drug on each organ system. Because of the potential contamination of the carotid ultrasound images-based predictor by the tissue surrounding the arteries, we recommend that other predictors, such as the pulse wave analysis-derived arterial age predictors and heart age predictors ^7, 8^, be used conjointly when assessing the effects of therapeutic interventions or lifestyle changes on the cardiovascular age.

## Methods

### Data and materials availability

We used the UK Biobank (project ID: 52887). The main code can be found at https://github.com/Deep-Learning-and-Aging. The code for survival prediction can be found at: The code for the survival analysis can be found at https://github.com/alanlegoallec/Deep_survival_from_time_series. The results can be interactively and extensively explored at https://www.multidimensionality-of-aging.net/. We will make the biological age phenotypes available through UK Biobank upon publication. The GWAS results can be found at https://www.dropbox.com/s/59e9ojl3wu8qie9/Multidimensionality_of_aging-GWAS_results.zip?d<u> l=0.</u>

### Software

Our code can be found at https://github.com/Deep-Learning-and-Aging. For the genetics analysis, we used the BOLT-LMM ^27, 28^ and BOLT-REML ^29^ softwares. We coded the parallel submission of the jobs in Bash ^30^.

### Cohort Dataset: Participants of the UK Biobank

We leveraged the UK Biobank^31^ cohort (project ID: 52887). The UKB cohort consists of data originating from a large biobank collected from 502,211 de-identified participants in the United Kingdom that were aged between 37 years and 74 years at enrollment (starting in 2006). The sample sizes for the different datasets leveraged in our study, after preprocessing, can be found in Figure 1A. The Harvard internal review board (IRB) deemed the research as non-human subjects research (IRB: IRB16-2145).

### Data types and Preprocessing

The data preprocessing step is different for the different data modalities: demographic variables, scalar predictors, time series and images. We define scalar predictors as predictors whose information can be encoded in a single number, such as blood pressure, as opposed to data with a higher number of dimensions such as time series (one dimension, which is time) and images (two dimensions, which are the height and the width of the image).

#### Demographic variables

First, we removed out the UKB samples for which age or sex was missing. For sex, we used the genetic sex when available, and the self-reported sex when genetic sex was not available. We computed age as the difference between the date when the patient attended the assessment center and the year and month of birth of the patient to estimate the patient’s age with greater precision. We one-hot encoded ethnicity.

#### Scalar data

We define scalar data as a variable that is encoded as a single number, such as blood pressure or pulse rate, as opposed to data with a higher number of dimensions, such as time series, and images. The complete list of scalar biomarkers can be found in Table S9 under “Arterial”. We did not preprocess the scalar data, aside from the normalization that is described under cross-validation further below.

#### Time series: pulse wave analysis

The UKB contains 234,837 samples with pulse wave analysis measurements, a measure of arterial stiffness (field 4205). Each sample is a time series of 100 measurements, collected over one second (Figure 1D). We scaled each sample independently between 0 and 1 and used the normalizing constant as a “side predictor” for the predictive algorithms. We name side predictors scalar predictors that can be fed to a neural network along with other data modalities (time series, images, videos) to improve the predictions. Here, the normalized pulse wave analysis measurements encode the information about the shape of the time series function (Volume=f(time)), while the side predictor encodes the information about the actual height of this function (the maximum value it takes). We did not filter out any samples.

We built simple LSTM and GRU architectures, as well. These architectures underperformed during our preliminary analysis (R^2^=14%) and we therefore did not train these models for our final pipeline.

#### Images: carotid ultrasound

The UKB contains carotid artery ultrasounds from both the left (field 20222, 8,992 images for 8,965 participants) and right (field 20223, 8,995 samples for 8,968 participants) sides. For each artery, four different measures are taken. One along the short axis, one along the long axis, one at an angle of 120 degrees and one at an angle of 150 degrees. The images are screenshots of the software used to record the ultrasound images (Fig. S11), so we extracted the actual images with a resulting final dimension of 506*437 pixels. We resized these images to 337*291 pixels. We superimposed the images taken along the long axis at different angles (long axis, 120 degree and 150 degrees) to obtain RGB images that we named “Mixed”. A sample of preprocessed carotid images can be found in Fig. S12.

#### Data augmentation for images

To prevent overfitting and increase our sample size during the training we used data augmentation ^32^ on the images. Each image was randomly shifted horizontally by up to 20%. We chose this hyperparameter for the transformations’ uniform distribution to represent the variations we observed between the images in the initial dataset.

The data augmentation process is dynamically performed during the training. Augmented images are not generated in advance. Instead, each image is randomly augmented before being fed to the neural network for each epoch during the training.

### Machine learning algorithms

For scalar datasets, we used elastic nets, gradient boosted machines [GBMs] and fully connected neural networks. For times series and images we used one-dimensional, two-dimensional, and three-dimensional convolutional neural networks, respectively.

#### Scalar data

We used three different algorithms to predict age from scalar data (non-dimensional variables, such as laboratory values). Elastic Nets [EN] (a regularized linear regression that represents a compromise between ridge regularization and LASSO regularization), Gradient Boosted Machines [GBM] (LightGBM implementation ^33^), and Neural Networks [NN]. The choice of these three algorithms represents a compromise between interpretability and performance. Linear regressions and their regularized forms (LASSO ^34^, ridge ^35^, elastic net ^36^) are highly interpretable using the regression coefficients but are poorly suited to leverage non-linear relationships or interactions between the features and therefore tend to underperform compared to the other algorithms. In contrast, neural networks ^37, 38^ are complex models, which are designed to capture non-linear relationships and interactions between the variables. However, tools to interpret them are limited ^39^ so they are closer to a “black box”. Tree-based methods such as random forests ^40^, gradient boosted machines ^41^ or XGBoost ^42^ represent a compromise between linear regressions and neural networks in terms of interpretability. They tend to perform similarly to neural networks when limited data is available, and the feature importances can still be used to identify which predictors played an important role in generating the predictions. However, unlike linear regression, feature importances are always non-negative values, so one cannot interpret whether a predictor is associated with older or younger age. We also performed preliminary analyses with other tree-based algorithms, such as random forests ^40^, vanilla gradient boosted machines ^41^ and XGBoost ^42^. We found that they performed similarly to LightGBM, so we only used this last algorithm as a representative for tree-based algorithms in our final calculations.

#### Time series: Pulse wave analysis

To analyse the pulse wave analysis data we built an eight layers deep convolutional neural network [CNN] whose architecture can be found in Supplementary Figure 68 and Supplementary Figure 69. The architecture consists of a convolutional block and a dense block. The inputs for the convolutional block are time series with a single channel and 100 time steps.

The convolutional block is composed of three convolutional layers with kernels of size three and strides of one. We used zero-padding so that the convolution would not reduce the spatial dimension of the input. The number of filters for these three convolutional layers are respectively 32, 64 and 128. We used one dimensional max pooling with a kernel size of 2 after each of the first two layers to reduce the dimension of the input, and we used a global max one-dimensional layer after the last convolutional layer to convert each of the 128 filters to a scalar feature. We then concatenated these 128 scalar features with the scaling factor of the imputed sample, duplicated 12 times (the scaling factor is a measure of the amplitude of the raw signal before it was normalized between zero and one, as described under Methods - Data types and Preprocessing - Time series - Pulse Wave Analysis). We inputted these 140 features into the dense block, which consists of two dense layers with respectively 128 and 64 nodes. We concatenated the 64 features outputted by second dense layer with sex and ethnicity. We inputted these 89 features into two dense layers with respectively 64 and 32 nodes. We used batch normalization, kernel regularization, bias regularization, ReLU activation for the seven first layers of the architecture. We used dropout to regularize the four dense layers, as well. Finally, we used a dense layer with a single node and linear activation as the final eighth layer to predict chronological age. We refer to the five dense layers as the dense block.

We built simple LSTM and GRU architectures, as well. These architectures underperformed during our preliminary analysis (R^2^=14%) and we therefore did not train these models for our final pipeline.

### Images

#### Convolutional Neural Networks Architectures

We used transfer learning ^43–45^ to leverage two different convolutional neural networks ^46^ [CNN] architectures pre-trained on the ImageNet dataset ^47–49^ and made available through the python Keras library ^50^: InceptionV3 ^51^ and InceptionResNetV2 ^52^. We considered other architectures such as VGG16 ^53^, VGG19 ^53^ and EfficientNetB7 ^54^, but found that they performed poorly and inconsistently on our datasets during our preliminary analysis and we therefore did not train them in the final pipeline. For each architecture, we removed the top layers initially used to predict the 1,000 different ImageNet images categories. We refer to this truncated model as the “base CNN architecture”.

We added to the base CNN architecture what we refer to as a “side neural network”. A side neural network is a single fully connected layer of 16 nodes, taking the sex and the ethnicity variables of the participant as input. The output of this small side neural network was concatenated to the output of the base CNN architecture described above. This architecture allowed the model to consider the features extracted by the base CNN architecture in the context of the sex and ethnicity variables. For example, the presence of the same anatomical arterial feature can be interpreted by the algorithm differently for a male and for a female. We added several sequential fully connected dense layers after the concatenation of the outputs of the CNN architecture and the side neural architecture. The number and size of these layers were set as hyperparameters. We used ReLU ^55^ as the activation function for the dense layers we added, and we regularized them with a combination of weight decay ^56, 57^ and dropout ^58^, both of which were also set as hyperparameters. Finally, we added a dense layer with a single node and linear activation to predict age.

#### Compiler

The compiler uses gradient descent ^59, 60^ to train the model. We treated the gradient descent optimizer, the initial learning rate and the batch size as hyperparameters. We used mean squared error [MSE] as the loss function, root mean squared error [RMSE] as the metric and we clipped the norm of the gradient so that it could not be higher than 1.0 ^61^.

We defined an epoch to be 32,768 images. If the training loss did not decrease for seven consecutive epochs, the learning rate was divided by two. This is theoretically redundant with the features of optimizers such as Adam, but we found that enforcing this manual decrease of the learning rate was sometimes beneficial. During training, after each image has been seen once by the model, the order of the images is shuffled. At the end of each epoch, if the validation performance improved, the model’s weights were saved.

We defined convergence as the absence of improvement on the validation loss for 15 consecutive epochs. This strategy is called early stopping ^62^ and is a form of regularization. We requested the GPUs on the supercomputer for ten hours. If a model did not converge within this time and improved its performance at least once during the ten hours period, another GPU was later requested to reiterate the training, starting from the model’s last best weights.

### Training, tuning and predictions

We split the entire dataset into ten data folds. We then tuned the models built on scalar data, on time series, on images and on videos using four different pipelines. For scalar data-based models, we performed a nested-cross validation. For time series-based and images-based, we manually tuned some of the hyperparameters before performing a simple cross-validation. We describe the splitting of the data into different folds and the tuning procedures in greater detail in the Supplementary.

### Interpretability of the machine learning predictions

To interpret the models, we used the regression coefficients for the elastic nets, the feature importances for the GBMs, a permutation test for the fully connected neural networks, and attention maps (saliency and Grad-RAM) for the convolutional neural networks (Supplementary Methods).

### Ensembling to improve prediction and define aging dimensions

We built a three-level hierarchy of ensemble models to improve prediction accuracies. At the lowest level, we combined the predictions from different algorithms on the same arterial subdimension. For example, we combined the predictions generated by the elastic net, the gradient boosted machine and the neural network on the pulse wave analysis scalar predictors (subdimension). At the second level, we combined the predictions from different subdimensions of a unique dimension. For example, we combined the pulse wave analysis subdimensions “Scalars” and “Time Series” into an ensemble prediction. Finally, at the highest level, we combined the predictions from the three arterial aging dimensions into a general arterial age prediction. The ensemble models from the lower levels are hierarchically used as components of the ensemble models of the higher models. For example, the ensemble models built by combining the algorithms at the lowest level for each of the carotid ultrasound subdimensions are leveraged when building the general arterial aging ensemble model.

We built each ensemble model separately on each of the ten data folds. For example, to build the ensemble model on the testing predictions of the data fold #1, we trained and tuned an elastic net on the validation predictions from the data fold #0 using a 10-folds inner cross-validation, as the validation predictions on fold #0 and the testing predictions on fold #1 are generated by the same model (see Methods - Training, tuning and predictions - Images - Scalar data - Nested cross-validation; Methods - Training, tuning and predictions - Images - Cross-validation). We used the same hyperparameters space and Bayesian hyperparameters optimization method as we did for the inner cross-validation we performed during the tuning of the non-ensemble models.

To summarize, the testing ensemble predictions are computed by concatenating the testing predictions generated by ten different elastic nets, each of which was trained and tuned using a 10-folds inner cross-validation on one validation data fold (10% of the full dataset) and tested on one testing fold. This is different from the inner-cross validation performed when training the non-ensemble models, which was performed on the “training+validation” data folds, so on 9 data folds (90% of the dataset).

### Evaluating the performance of models

We evaluated the performance of the models using two different metrics: R-Squared [R^2^] and root mean squared error [RMSE]. We computed a confidence interval on the performance metrics in two different ways. First, we computed the standard deviation between the different data folds. The test predictions on each of the ten data folds are generated by ten different models, so this measure of standard deviation captures both model variability and the variability in prediction accuracy between samples. Second, we computed the standard deviation by bootstrapping the computation of the performance metrics 1,000 times. This second measure of variation does not capture model variability but evaluates the variance in the prediction accuracy between samples.

### Arterial age definition

We defined the biological age of participants as the prediction generated by the model corresponding to arterial dimension or subdimension, after correcting for the bias in the residuals.

We indeed observed a bias in the residuals. For each model, participants on the older end of the chronological age distribution tend to be predicted younger than they are. Symmetrically, participants on the younger end of the chronological age distribution tend to be predicted older than they are. This bias does not seem to be biologically driven. Rather it seems to be statistically driven, as the same 60-year-old individual will tend to be predicted younger in a cohort with an age range of 60-80 years, and to be predicted older in a cohort with an age range of 60-80. We ran a linear regression on the residuals as a function of age for each model and used it to correct each prediction for this statistical bias.

After defining biological age as the corrected prediction, we defined accelerated aging as the corrected residuals. For example, a 60-year-old whose carotid ultrasound data predicted an age of 70 years old after correction for the bias in the residuals is estimated to have an arterial age of 70 years, and an accelerated arterial aging of ten years.

It is important to understand that this step of correction of the predictions and the residuals takes place after the evaluation of the performance of the models but precedes the analysis of the biological ages properties.

### Genome-wide association of accelerated arterial aging

The UKB contains genome-wide genetic data for 488,251 of the 502,492 participants^63^ under the hg19/GRCh37 build.

We used the average accelerated aging value over the different samples collected over time (see Supplementary - Generating average predictions for each participant). Next, we performed genome wide association studies [GWASs] to identify single-nucleotide polymorphisms [SNPs] associated with arterial accelerated agings (general, carotid ultrasound-measured, and pulse wave analysis-measured) using BOLT-LMM ^27, 28^ and estimated the the SNP-based heritability for each of our biological age phenotypes, and we computed the genetic pairwise correlations between dimensions using BOLT-REML ^29^. We used the v3 imputed genetic data to increase the power of the GWAS, and we corrected all of them for the following covariates: age, sex, ethnicity, the assessment center that the participant attended when their DNA was collected, and the 20 genetic principal components precomputed by the UKB. We used the linkage disequilibrium [LD] scores from the 1,000 Human Genomes Project ^64^. To avoid population stratification, we performed our GWAS on individuals with White ethnicity.

#### Identification of SNPs associated with accelerated aging

We identified the SNPs associated with accelerated arterial aging dimensions and subdimensions using the BOLT-LMM ^27, 28^ software (p-value of 5e-8). The sample size for the genotyping of the X chromosome is one thousand samples smaller than for the autosomal chromosomes. We therefore performed two GWASs for each aging dimension. (1) excluding the X chromosome, to leverage the full autosomal sample size when identifying the SNPs on the autosome. (2) including the X chromosome, to identify the SNPs on this sex chromosome. We then concatenated the results from the two GWASs to cover the entire genome, at the exception of the Y chromosome.

We plotted the results using a Manhattan plot and a volcano plot. We used the bioinfokit ^65^ python package to generate the Manhattan plots. We generated quantile-quantile plots [Q-Q plots] to estimate the p-value inflation as well.

#### Heritability and genetic correlation

We estimated the heritability of the accelerated aging dimensions using the BOLT-REML ^29^ software. We included the X chromosome in the analysis and corrected for the same covariates as we did for the GWAS. Using the same software and parameters, we computed the genetic correlations between carotid ultrasound-based and pulse wave analysis-based accelerated arterial aging.

We annotated the significant SNPs with their matching genes using the following four steps pipeline. (1) We annotated the SNPs based on the rs number using SNPnexus ^66–70^. When the SNP was between two genes, we annotated it with the nearest gene. (2) We used SNPnexus to annotate the SNPs that did not match during the first step, this time using their genomic coordinates. After these two first steps, 30 out of the 9,697 significant SNPs did not find a match. (3) We annotated these SNPs using LocusZoom ^71^. Unlike SNPnexus, LocusZoom does not provide the gene types, so we filled this information with GeneCards ^72^. After this third step, four genes were not matched. (4) We used RCSB Protein Data Bank ^73^ to annotate three of the four missing genes. One gene on the X chromosome did not find a match (position 56,640,134).

### Non-genetic correlates of accelerated aging

We identified non-genetically measured (i.e factors not measured on a GWAS array) correlates of each aging dimension, which we classified in six categories: biomarkers, clinical phenotypes, diseases, family history, environmental, and socioeconomic variables. We refer to the union of these association analyses as an X-Wide Association Study [XWAS]. (1) We define as biomarkers the scalar variables measured on the participant, which we initially leveraged to predict age (e.g. blood pressure, Table S9). (2) We define clinical phenotypes as other biological factors not directly measured on the participant, but instead collected by the questionnaire, and which we did not use to predict chronological age. For example, one of the clinical phenotypes categories is eyesight, which contains variables such as “wears glasses or contact lenses”, which is different from the direct refractive error measurements performed on the patients, which are considered “biomarkers” (Table S12). (3) Diseases include the different medical diagnoses categories listed by UKB (Table S15). (4) Family history variables include illnesses of family members (Table S18). (5) Environmental variables include alcohol, diet, electronic devices, medication, sun exposure, early life factors, medication, sun exposure, sleep, smoking, and physical activity variables collected from the questionnaire (Table S21). (6) Socioeconomic variables include education, employment, household, social support and other sociodemographics (Table S24). We provide information about the preprocessing of the XWAS in the Supplementary Methods.

### Prediction of survival from pulse wave analysis records

The UKB cohort also contains mortality data for 30,203 participants, 6,000 of which had their pulse wave recorded. We predicted survival using different Cox Proportional Hazard [CoxPH] models ^74^. An elastic net, a gradient boosted machine [GBM], a fully connected neural network, and a one-dimensional convolutional neural network [CNN] with a side, fully connected neural network for scalar side inputs (Fig. S13). We used scikit-survival ^75^ library’s CoxPHFitter (elastic net) and GradientBoostingSurvivalAnalysis (GBM) functions, PyTorch ^76^ and torchtuples’ libraries MLPVanilla function (neural network), and we built the CNN architectures using TensorFlow-v2. We adapted the loss function for the CNN survival model from a tutorial by Sebastian Pölsterl: https://github.com/sebp/survival-cnn-estimator/blob/master/tutorial tf2.ipynb.

For the elastic nets, we tuned the regularization strength (loguniform distribution from 1e-10 to 1) and the ratio between L1 and L2 regularization (uniform distribution between 0 and 1). For the GBM, we tuned the maximum number of tree leaves for a single tree (integer uniformly sampled from 5 to 45), the minimum number of samples per leaf (integer uniform from 100 to 500), the minimum number of hessian weight per leaf (integer uniform from -5 to 4), the subsample ratio of samples for each tree (uniform from 0.2 to 1.0), the subsample ratio of features considered for each tree (uniform from 0.4 to 1.0), the L1 regularization (loguniform from 1e-2 to 1e2), the L2 regularization (loguniform from 1e-2 to 1e2) and the number of trees included in the model (inte-ger uniform from 150 to 450). For the fully connected neural networks, we tuned the initial learning rate (loguniform distribution from 1e-5 to 0.1) and the weight decay (loguniform distribution from 1e-6 to 1e3). For the CNNs, we tuned the learning rate (loguniform distribution from 1e-5 to 1e-3), the weight decay (loguniform distribution from 1e-5 to 1e3), the dropout rate (uniform from 0 to 0.9), the presence of batch normalization after convolutional layers (choice: True/False) and the presence of batch normalization after dense layers (choice: True/False).

The code for the survival analysis can be found at https://github.com/alanlegoallec/Deep_survival_from_time_series.

## Data Availability

We used the UK Biobank (project ID: 52887). The main code can be found at https://github.com/Deep-Learning-and-Aging. The code for survival prediction can be found at: The code for the survival analysis can be found at https://github.com/alanlegoallec/Deep_survival_from_time_series. The results can be interactively and extensively explored at https://www.multidimensionality-of-aging.net/. We will make the biological age phenotypes available through UK Biobank upon publication. The GWAS results can be found at https://www.dropbox.com/s/59e9ojl3wu8qie9/Multidimensionality_of_aging-GWAS_results.zip?dl=0.

https://github.com/alanlegoallec/Deep_survival_from_time_series

https://www.multidimensionality-of-aging.net/

https://www.dropbox.com/s/59e9ojl3wu8qie9/Multidimensionality_of_aging-GWAS_results.zip?dl=0

## Acknowledgments

We would like to thank Raffaele Potami from Harvard Medical School research computing group for helping us utilize O2’s computing resources. We thank HMS RC for computing support. We also want to acknowledge UK Biobank for providing us with access to the data they collected. The UK Biobank project number is 52887.

## Author Contributions

**Alan Le Goallec:** (1) Designed the project. (2) Supervised the project. (3) Preprocessed the pulse wave analysis records. (4) Predicted chronological age from carotid ultrasound images. (5) Computed the attention maps for carotid ultrasound images. (6) Ensembled the models, evaluated their performance, computed arterial ages and estimated the correlation structure between the arterial aging dimensions. (7) Performed the genome wide association studies. (8) Performed the survival analysis. (9) Designed the website. (10) Wrote the manuscript.

**Sasha Collin:** (1) Preprocessed the carotid ultrasound images. (2) Predicted chronological age from the pulse wave analysis records. (3) Computed the attention maps for the pulse wave analysis records.

**Samuel Diai:** (1) Predicted chronological age from scalar features. (2) Wrote the python class to build an ensemble model using a cross-validated elastic net. (3) Performed the X-wide association study. (4) Implemented a first version of the website https://www.multidimensionality-of-aging.net/.

**Théo Vincent:** (1) Website data engineer. (2) Implemented a second version of the website https://www.multidimensionality-of-aging.net/.

**Chirag J. Patel:** (1) Supervised the project. (2) Edited the manuscript. (3) Provided funding.

